# Protocol for the Development of Artificial Intelligence Models for the Reduction of Surgical Complications Based on Intraoperative Video - Surg_Cloud project

**DOI:** 10.1101/2024.05.26.24307908

**Authors:** Antonio Sampaio Soares, Sophia Bano, Laura T Castro, Ricardo Rocha, Paulo Alves, Paulo Mira, João Costa, Manish Chand, Danail Stoyanov

## Abstract

**Introduction:** Complications following abdominal surgery have a very significant negative impact on the patient and the health care system. Despite the spread of minimally invasive surgery, there is no automated way to use intraoperative video to predict complications. New developments in data storage capacity and artificial intelligence algorithm creation now allow for this opportunity.

**Methods:** Development of deep learning algorithms through supervised learning based on the Clavien-Dindo scale to categorise postoperative outcomes in minimally invasive abdominal surgery. An open-source dataset will be built, which will not only include intraoperative variables but also data related to patient outcomes, making it more generalisable and useful to the scientific community. This dataset will be shared under a non-commercial use license to promote scientific collaboration and innovation.

**Expected Results:** The planned outputs include the publication of a research protocol, main results, and the open-source dataset. Through this initiative, the project seeks to significantly advance the field of artificial intelligence-assisted surgery, contributing to safer and more effective practice.

## 1 Introduction

The conventional approach to surgical training is deeply rooted in a model that combines theoretical teaching with supervised practice in the operating environment. However, this model has its flaws. One of the most notable is the difficulty in predicting the variability that occurs during surgery itself, whether due to each patient’s unique anatomy or the specific pathology that justifies the intervention. Although surgeons accumulate a wealth of information during each procedure, this data often remains confined within the walls of the operating room. This is largely due to the lack of efficient mechanisms for capturing this data and the subsequent inability to analyse it systematically and rigorously. This situation creates a vicious cycle where learning becomes fragmented and inefficient, negatively affecting clinical outcomes and the evolution of surgical practice.

The emergence of minimally invasive surgery marked a revolution in clinical practice. In addition to minimising trauma and accelerating patient recovery, this approach provides an unprecedented opportunity: the ability to record the entire surgical procedure on video. Previous works, such as those by Birkmeyer [1] and Curtis [2], have already provided valuable insights by demonstrating that the quality of surgical gestures is correlated with clinical outcomes. These works were based on a still very rudimentary human assessment that cannot be scaled. With the advent of new technologies in image analysis and artificial intelligence (AI), it is now possible to automate the evaluation of these videos. The objective of this project is to go beyond simple evaluation, developing AI algorithms that can provide real-time support during the surgical act.

Post-surgical complications, both in minimally invasive cholecystectomy and colectomy, have a very high human and economic cost. They increase patient mortality and morbidity and burden health systems with additional costs in postoperative care and prolonged hospital stays. In this context, it is not only desirable, but ethically imperative, to seek new ways to reduce the risk of complications and improve clinical outcomes. The use of AI algorithms for intraoperative video analysis has the potential to revolutionise how safety in surgery is approached, allowing for the identification and correction of problems in real time and thus significantly improving the quality of care provided.

## 2 Methods

### 2.1 Dataset construction

#### 2.1.1 Dataset summary

This dataset will cover both preoperative data, intraoperative data and video and 30 day postoperative outcomes on two groups of patients regularly operated in a minimally invasive fashion. These are patients undergoing cholecystectomy and colorectal resection. Video data will be collected from the minimally invasive device monitor (laparoscopic or robotic). Other variables will be tabular data.

#### 2.1.2 Dataset identity and access

The dataset is going to be called Surg_Cloud Cholecystectomy and Surg_Cloud Colorectal, including the version released (V1, V2, …). The first release is expected in the first semester of 2025. It will be shared through a web host with login credentials available for researchers. The dataset will be made available under a CC BY-NC-SA license. This allows its use for research purposes but restricts its use for commercial purposes. This balance aims to promote scientific collaboration and innovation while protecting the data from unauthorised commercial exploitation.

#### 2.1.3 Reasons behind dataset creation and its purpose(s)

This dataset was created to increase the data available for research in Artificial Intelligence in surgery. This data will be collected on a number of patients reflective of day to day practice. It will include outcome data to allow the use of this knowledge to guide the study of the surgical procedure. The intended benefit is to allow the development of technological solutions to improve patient care. Different teams can obtain different insights from their analysis of the data, which is why the aim of the study group is to create an open source dataset.

#### 2.1.4 Data origin

Data is collected from patients receiving healthcare under normal circumstances. No change in normal clinical practice will occur. Every patient approached will be told the aims of this study, the absence of change in routine clinical care and that only completely anonymised data will be made public in a dataset. This information is specified on the informed consent form every patient signs. The consent form has been approved in every participating hospital.

#### 2.1.5 Data sampling and aggregation from multiple sources

The dataset will be composed of anonymised patient data from multiple hospitals. The enrolment of patients will occur under day to day clinical conditions, reflecting a convenience sample that does not necessarily reflect clinical practice. It is to be expected that sicker patients will not be as represented in the study participants as more clinically stable ones, given the context of the provision of care.

#### 2.1.6 Data shifts over time

Every dataset released will have a corresponding version number where the details of data acquisition will be made explicit. If any significant changes occur between versions of the release, these will also be detailed in the accompanying documentation. It is expected that most variation will occur in the setting of new patients being added.

#### 2.1.7 Composition of groups within the dataset

Patients will be grouped according to standard practice as mentioned in the data collection forms, found in tables 1 and 2. Besides the video and the data mentioned in this protocol, no other data will be collected from patients.

#### 2.1.8 Recording of Individuals’ Attributes

Patient level data will be composed of the variables contained in the tables found in the end, 1 and2. This data will be available for every patient.

#### 2.1.9 Groups at risk of disparate health outcomes

The dataset comprises various attributes that reflect patient interactions in public hospitals. These attributes were selected to provide insights into common healthcare practices and patient outcomes in these settings. Each attribute is presented at an individual level, ensuring a granular and detailed analysis. However, it’s important to note that no patient-identifiable data has been collected to preserve privacy.

#### 2.1.10 Limitations of the dataset

The dataset will be created from videos collected in very diverse range of hospitals. The patient selection will be mostly a convenience sample, given the constraints of surgical care provision. Data quality will be variable given the range of devices that collect the data. These specifics will be detailed on a per patient basis.

#### 2.1.11 Modifications made to the data

The only data modifications planned are the blurring of out of body images to ensure complete anonymisation. No data on the hospital where the data was collected will be shared. No further modifications are planned at this stage. No synthetic data will be used.

#### 2.1.12 Missing data

No missing data will be imputed.

#### 2.1.13 Known or potential bias caused or exacerbated by data acquisition and processing

Given the complete anonymisation of patients and the lack of impact in the patient’s clinical care, no bias is expected to be exacerbated by the creation of this dataset.

#### 2.1.14 Known or potential exclusion introduced by data collection

Formal exclusion criteria have not been defined. Patients less likely to be included in the dataset are urgent operations where the patients is likely sicker. This will necessarily impact on the ability of the team to ensure proper informed consent. Due to this fact, a lesser number of urgent patients are expected to be included.

#### 2.1.15 Known or potential bias in assigned or derived labels

Labels will be assigned as per the data on tables 1 and 2. Variables included are well standardised. Postoperative complications will be categorised using the Clavien-Dindo scale[3], which has 5 levels of severity, from I (minor deviations from the desired postoperative course) to V (patient death). This scale provides a structured framework for classifying and comparing complications, allowing for more precise analysis.

#### 2.1.16 Ethics and governance

Patient inclusion will necessitate ethical approval at every hospital and informed consent from every patient recruited. No patient identifiable data will be shared in the dataset. The study team will have a steering committee, hospital leads and collaborators. Collaborative authorship guidelines will be followed. The study has already received approval by the Hospital Prof. Doutor Fernando Fonseca Ethics Committee, approval number 113 / 2023.

#### 2.1.17 Patient and public participation

A formal patient and public involvement initiative is planned to be developed before dataset publication.

#### 2.1.18 Importance of Reproducibility and Generalisability

Reproducibility of results in research is crucial for the credibility and advancement of science. In accordance with FAIR principles (Findable, Accessible, Interoperable, and Reusable) [4], this project aims not only to conduct rigorous studies but also to make data and methods available in a way that allows for their verification and reuse by other researchers. Dataset development was made according to the STANDING recommendations [5]. By doing so, we increase the likelihood that the results are generalizable and, consequently, more useful globally. The creation of an open dataset follows the example of successful initiatives such as the Heidelberg Colorectal Dataset, published in the journal Nature [6]. This dataset is a reference in the field, consisting of 30 videos of colorectal surgeries, accompanied by annotations and extensive documentation. Other institutions, such as IRCAD in the case of laparoscopic cholecystectomy [7], and the University of Dresden [8], have also contributed to science by publishing open-source datasets.

### 2.2 Analysis plan

#### 2.2.1 Objetives

Development of a convolutional neural network with supervised methodology for predicting the grade of postoperative complication using the Clavien-Dindo scale.

#### 2.2.2 Study design

Multicentric prospective cohort study for patients undergoing minimally invasive cholecystectomy or colorectal resection.

#### 2.2.3 Descriptive analysis

In the initial phase, a descriptive statistical analysis of demographic, intraoperative, and postoperative variables will be conducted. This analysis aims to provide a quantitative summary of the data, helping to define the profile of the patients and the surgical interventions performed. Measures of central tendency, dispersion, and distribution will be calculated for each variable, such as mean, standard deviation, median, and percentiles.

#### 2.2.4 Regression analysis

Subsequently, a regression analysis will be conducted to identify variables that may predict postoperative complications. This regression model will be adjusted to account for potential confounding factors and will allow the assessment of the independent impact of each predictor variable. The analysis will be conducted for both categorical and quantitative variables.

#### 2.2.5 Statistical Significance and Interpretation of Results

All statistical tests will be performed with a pre-defined significance level of p = 0.05, and the results will be interpreted in the light of the clinical context to ensure that the conclusions are not only statistically significant but also clinically relevant.

### 2.3 Algorithm development

#### 2.3.1 Compliance with methodological guidelines

The development of AI algorithms for this project will be rigorously guided by best practices. The TRIPOD AI [9] guidance will be followed. These standards ensure that the development, validation, and implementation of AI algorithms in medical contexts follow strict standards to ensure their efficacy and safety. The publication of STARD AI [10] is expected soon, and according to the timeline of this project, its use in publications originating from it is anticipated.

#### 2.3.2 Supervised learning approach

The algorithms will be trained using a supervised learning approach, with the categories of the Clavien-Dindo scale serving as the outcome variable or ‘label’. Supervised learning was chosen to allow more focused and accurate training, considering the specific and complex characteristics of the medical data involved.

The collected video data will be divided into three sets: training, validation, and testing. The proportion for this division will be 70%, 15%, and 15%, respectively. This structure aims to ensure that the model is trained on a large amount of data while reserving an adequate portion for validation and testing, thereby minimising the risk of overfitting.

Python programming language, widely recognised for its versatility and robustness in AI projects, will be used. The specific infrastructure for training the neural networks will be PyTorch, an open-source machine learning library that is highly effective for implementing convolutional neural networks.

The specific type of neural network to be used will be the Convolutional Neural Network (CNN), which is currently considered the ‘state-of-the-art’ methodology for the analysis of medical images. CNNs have demonstrated an exceptional ability to identify patterns in images, making them ideal for the task at hand.

#### 2.3.3 Evaluation and continual improvement

After initial training, the algorithm will undergo several rounds of evaluation and adjustment. Metrics such as accuracy, sensitivity, and specificity will be used to assess the model’s performance. The results of these evaluations will inform subsequent iterations of the algorithm, allowing for continuous improvement of its performance. Thus, the project aims to develop a highly effective and reliable AI algorithm that can be used to significantly improve surgical practice and patient outcomes.

### 2.4 Data storage

Data will be uploaded to a secure cloud storage service. This service will include encrypted storage and access controlled by verified credentials, ensuring that data access is controlled and traceable. Additionally, the platform allows for the connection with applications to run the code for training and testing the algorithm. Both clinical and video data are uploaded after a period of 30 days post-surgery, ensuring complete anonymisation of the uploaded data.

### 2.5 Collaborative authorship

Collaborative authorship will be attributed to members of the study group according to the established guidelines[11]. One hospital lead and up to three local collaborators will be eligible per centre.

## 3 Expected Results and Evidence of Feasibility

The development of AI algorithms is a rapidly growing and innovative area. Various fields, including healthcare, are beginning to feel the transformative impacts of these technologies. However, the implementation of AI in healthcare faces unique challenges, namely restrictions on access to high-quality data and the complexities associated with the high inherent risk in healthcare. Despite these challenges, there are already AI algorithms that have received regulatory approval, demonstrating that it is possible to meet the stringent demands of this field. This fact underlines the feasibility of developing and implementing AI algorithms in clinical environments.

In surgery, the main challenge is the lack of specific data related to patient outcomes. Many of the datasets currently available focus on intraoperative variables but neglect postoperative outcomes. Moreover, the data often come from tertiary academic institutions and involve highly selected cases, which limits their applicability in the real world.

This project aims to address these gaps by using data that more faithfully reflect the clinical reality experienced in diverse contexts. The goal is to develop an AI algorithm that can not only assist during the surgical procedure but also accurately predict postoperative complications.

## 4 Timeline

Centre recruitment is ongoing. Patient recruitment is expected to occur throughout 2024. Early algorithm development should start in the second semester 2024 with the first outputs expected in the first semester of 2025.

## Data Availability

All data produced in the present study will be available upon request to the authors

## A Cholecystectomy - data collection form

**Table 1.**
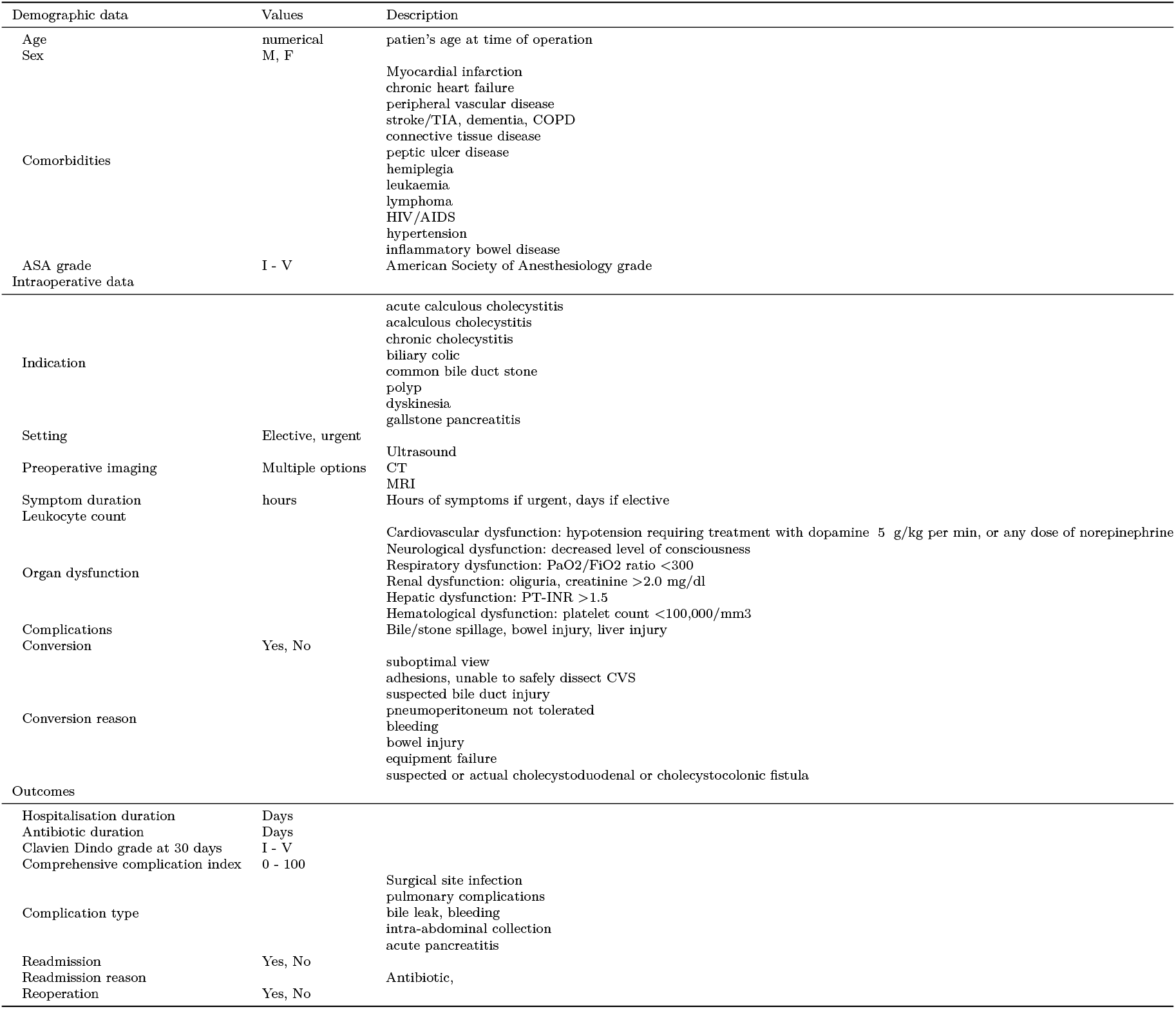
Case report form for cholecystectomy patients.

## B Colorectal resection and appendicectomy - data collection form

**Table 2.**
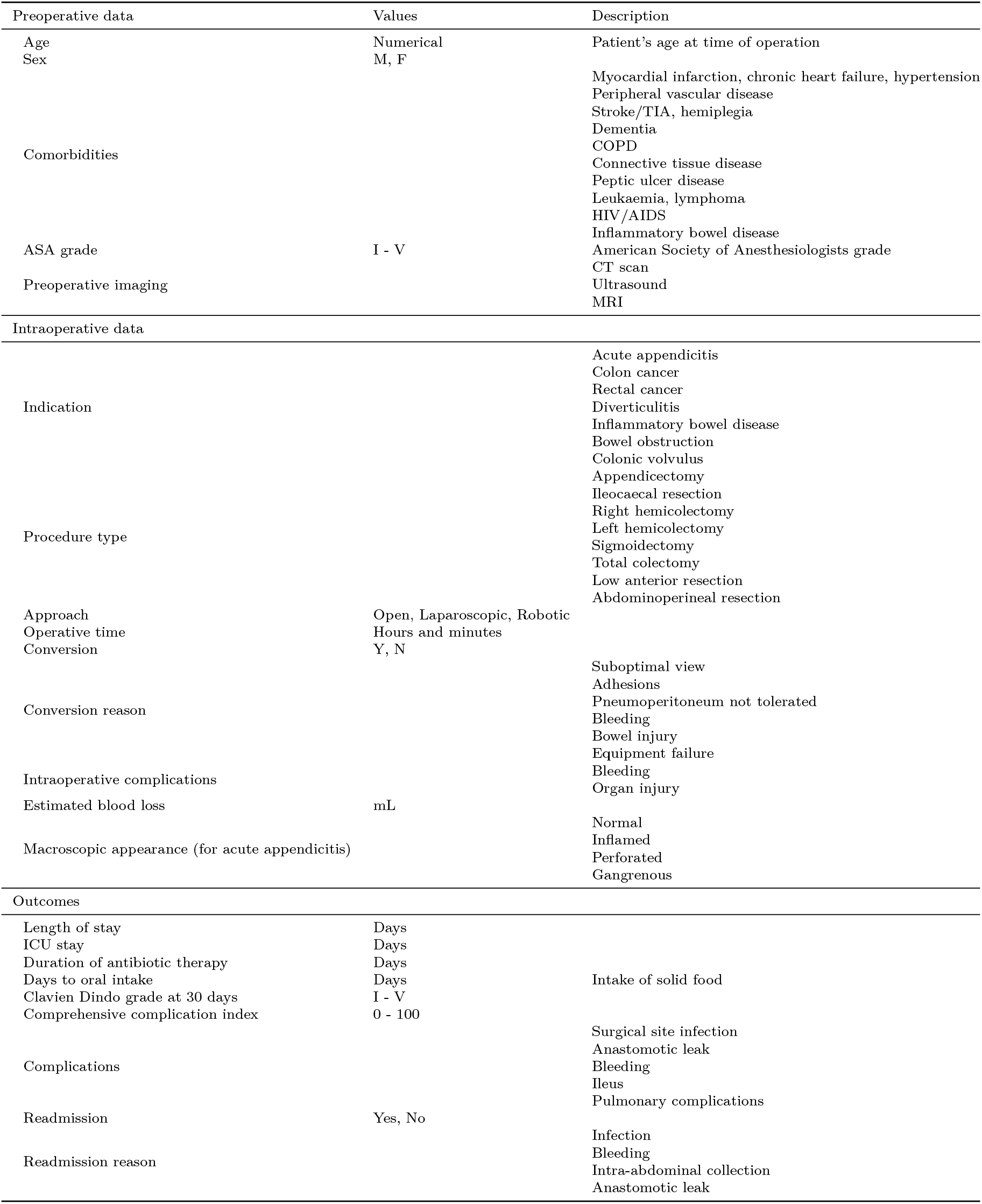
Case report form for colorectal resection patients.

